# “Comparison between topical Platelet Rich Plasma and Normal Saline dressing, in conjunction with Total Contact Casting in Treatment of Diabetic Foot Ulcer– a Randomized Control Trial”

**DOI:** 10.1101/2024.03.20.24304330

**Authors:** Subha Das, Sanyal Kumar, Sanjay Kumar Pandey, Anjani Kumar, Niraj Kumar, Ranjeet Kumar

**Author notes:** Corresponding Dr.Sanjay Kumar Pandey (Professor & Head), Department of PM&R, All India Institute of Medical Sciences, Phulwari Sharif, Patna-801507, India, E-mail ID.

## Abstract

**Introduction:** Wound care, which includes cleaning and fostering a moist wound healing environment, play a crucial role in the management of diabetic foot ulcers.. Total-contact casts are widely used as the most effective external technique for off-loading plantar ulcers. This study compared the effectiveness of PRP and normal saline dressings in conjunction with TCC.

**Methods:** 36 patients with diabetic foot ulcers were divided into two groups using computer generated randomization into PRP and NS groups with 18 patients in each group. The PRP group was given autologous PRP, and the NS group was given wet dressing with NS, following which TCC was applied in each case for off-loading. Follow-up was performed every 15 days for 90 days. In each follow-up measurement and TCC application, the time to heal and PUSH score was used to measure the condition of the wound.

**Results:** In the PRP group, the mean wound size 8.28±1.18 and the mean Push tool total 13.44±0.98 at base line which gradually decreased, and finally, on day 90, it was reduced to 0.61±1.20 and 1.89±3.68, respectively. In NS group the mean wound size 8.45±1.13 and mean Push tool total 13.50±0.92 at baseline which was gradually decreased and finally at day 90 it was reduced to 1.58±1.55 and 4.61±4.37 respectively. A significant difference was observed in both groups during the final evaluation. Compared with the NS group, the PRP group showed greater improvement.

**Conclusion:** Both autologous PRP and NS are effective in treating DFU, when used along with TCC. However, PRP therapy is better at reducing the healing time and hospital visits.

## Introduction

The metabolic condition known as Type II diabetes mellitus (DM2) is characterized by hyperglycemia, followed by insulin resistance [1]. Numerous co-morbidities, including peripheral neuropathy, chronic renal failure, stroke, cardiovascular disease, and diabetic foot ulcers, are linked to type 2 diabetes mellitus [2]. The most common side effect of type 2 DM is diabetic foot ulcers (DFUs), which can result in repeated hospital visits, hospital stays, and financial strain for the patients [3]. Poor blood glucose management and high plantar pressure are the primary causes of DFUs, which are defined as full-thickness wounds that penetrate the dermis (deep vascular and collagenous inner skin layer) and are located below the ankle in diabetic patients [4,5]. Diabetic ulcers present as excruciating ulcers that break down dermal tissues, including the dermis, epidermis, and subcutaneous tissue. The non-healing phenotype of diabetic foot ulceration has been attributed to immune system dysfunction, microbial invasion, and epithelial disintegration. 25% of patients with diabetes have a life-time risk of developing diabetic foot ulcers, the majority of whom will require amputation within four years of the initial diagnosis [3].

Owing to their multi-factorial etiology, foot ulcers are difficult to treat and place a significant burden on patients, health-care systems, and society [6]. Prevention is the key to managing diabetic foot wounds [7]. Patient education, regular foot examinations, and therapeutic foot-wear are essential to prevent DFU [7, 8]. Good clinical care entails frequent debridement, off-loading via casting with a walking rod, total contact cast (TCC), orthotic support, moist wound care with various dressing materials such as normal saline (NS), an antimicrobial agent, alginate, treatment of infection, and revascularization of the ischemic limb [9]. In severe wounds, however, vascular repair or amputation may be necessary [10, 11].15% of diabetic patients worldwide develop diabetic foot ulcers (DFU) at some point in their lives. An amputation of approximately 28% may be necessary [12]. The global prevalence of DFU is 6.3% in the overall population and is expected to increase in the future [13]. In India, the prevalence of diabetic foot ulcers is 4-5%, which is much lower than that reported in western countries [14].

## Materials and Methods

The study was conducted at a tertiary care center with a well-equipped research laboratory and other facilities for patient care after obtaining institutional ethical committee approval via AIIMS/Pat/IEC/PGTh/ July20/02 on September 29. The CONSORT statement requirements for reporting randomized controlled trials were followed, and the study complied with the principles of the Declaration of Helsinki. The clinical trial registration was performed (CTRI/2022/08/044973).

## Study design and sample

This randomized clinical trial was conducted in the Department of Physical Medicine and Rehabilitation, All India Institute of Medical Science, Patna, India. The study population was estimated using statistical formulas, and 36 patients were included in each group based on sample size calculations. Convenient sampling was used to recruit cases after fulfilling the inclusion and exclusion criteria from October 2021 to November 2022. Written informed consent was obtained from all the patients. Each participant underwent a complete examination. Patients were randomly assigned to two groups. The patients in the first group were administered PRP therapy along with TCC, where as the patients in the second group were administered NS dressing along with TCC. In this study, data will be collected, and the outcome will be measured in terms of time to heal and the PUSH Tool 3.0. The Statistical Package for the Social Sciences (SPSS) version 25 software will be used for statistical analysis after the collection of all relevant data.

## Study Procedure

### PRP preparation and application

Twenty milliliters of blood, taken from the basilic antecubital veins, was placed into vacuum-sterilized test tubes together with 3.8 milliliters of sodium citrate to serve as an anticoagulant. Using an Eppendorf Centrifuge 5720, the blood samples were centrifuged for 12 min at 1,200 rpm. Three layers of blood were isolated: red blood cells were found in the bottom layer, platelets and white blood cells were found in the thin middle layer, and white blood cells (the buffy coat) were found in the upper layer. The upper and intermediate buffy layers were then transferred to an empty sterile tube. To facilitate the development of soft pellets (erythrocytes and platelets) at the bottom of the tube, the plasma was centrifuged once more for seven minutes at 3,300 rpm; the upper two thirds of the platelet-poor plasma was discarded; pellets were homogenized in the lowest third (3mL) of the plasma to generate the PRP; the PRP was then extracted using a 5 ml syringe [15]. After measuring the wound, proper cleaning and debridement were performed. Freshly prepared autologous PRP was then applied to the ulcer bed. The wound was covered with sterile, dry, gauze. After proper covering, a total contact cast was applied for offloading ( Image1).

## NS dressing application

First the wound was measured. The wound was cleaned, debridement was performed, and normal saline-soaked gauze was applied over the ulcer bed. The wound was covered with sterile, dry gauze. After proper covering, a total contact cast was applied for offloading.

## TCC application

Inter-digital padding was applied first. Next, the stockinet was placed from the knee to the toes in a manner that showed the toes and prevented wrinkles or bunching. Layer of felt (D-filler) padding measuring 10-15 mm was positioned under the medial longitudinal arch. Cut to size, 5 mm silicone strips were placed across the bony prominences (e.g., medial/lateral malleoli, anterior tibia) [16]. Over the cushioning, a plaster of Paris cast was placed, extending one inch distal to the fibular head from the tips of the toes. To provide optimal contact, the cast was precisely shaped to fit the leg and foot [17, 18]. Patients were instructed to walk no more than one-third of their normal ambulation (Image 2).

## Follow-up and statistical analysis plan

Data collection (by“timetoheal”and“PUSHTool3.0”) will be performed at baseline and in the subsequent, 2 weekly follow-up, up to 3 months. In each follow-up measurement, debridement and TCC re-application were performed. The data were examined using IBM Corporation, Armonk; NYs, SPSS programme (version23). A significance threshold of p < 0.05 was established; the X 2 test was employed to compare qualitative factors, while the Unpaired T-test was ued to assess quantitative data. The standard deviation of the normal distribution of the quantitative variables was used to determine the significance level, which was set at p < 0.05.

## Results

In the PRP group, 11 (61.1%) patients were male and the remaining seven (38.9%) were female; in the NS group, 10(55.6%) were males and the remaining eight (44.4%) were females. Hence, in this study, 21 (58.3%) patients were males and 15 (41.7%) were females. The proportions of men and women in each group did not differ significantly (p = 0.735). Hence, the groups were matched according to their sex. The left and right sides were involved in equal proportions in both PRP and NS groups. The mean age of the PRP group was 56.50±4.96 years, while that of the NS group was 54.89±4.51 years. No significant difference was found in the mean age between the PRP and NS groups (p = 0.315). The mean BMI of the PRP group was 26.07±3.70 kg/m2, while the mean BMI of the NS group was 25.17±3.28 kg/m2. The PRP and NS groups mean BMIs did no significant differences (p=0.447) (Figure 1). The mean duration of DM of PRP group was 14.22±5.37 years while the mean duration of DM of NS group was 11.00±4.39 years. No significant difference was found in the mean duration between the PRP and NS group (p=0.057). The mean ABI of the PRP group was 0.99±0.12, while the mean ABI of the NS group was 0.98±0.15. No significant difference was found in the mean ABI between the PRP and NS group (p=0.811). The mean HbA1c level of the PRP group was 8.57±0.67 while the mean HbA1c of the NS group was 8.15±0.52. No significant difference was found in the mean HbA1c level between the PRP and NS group (p=0.53). The mean RBS of the PRP group was 217.22±27.78 while the mean HbA1c of the NS group was 208.00±23.51. No significant difference was found in the mean RBS between the PRP and NS group (p=0.290). The mean platelet count of the PRP group was 258.44±45.30, while that of the NS group was 243.22±42.32. No significant difference was found in the mean platelet count between the PRP and NS group (p=0.305). The mean albumin level in the PRP group was 3.77±0.38, while that in the NS group was 3.64±0.72. No significant difference was found in the mean platelet count between the PRP and NS group (p=0.508). The mean time to heal in PRP group was 11.17±2.73 weeks while the mean time to heal in NS group was 13.78±1.66 weeks. A significant difference was found in the mean time to healing between the PRP and NS group (Table1). In PRP group the mean wound size at baseline was 8.28±1.18 which was gradually decreased and finally at day 90 it was reduced to 0.61±1.20. In NS group the mean wound size at baseline was 8.45±1.13 which was gradually decreased and finally at day 90 it was reduced to 1.58±1.55. The PRP and NS groups showed a significant difference in mean wound size at subsequent follow up period (Table 2). From baseline to every follow up intervals the mean change in wound size in the PRP and NS groups was significant (p<0.001) (Table3). In PRP group the mean Push tool total at baseline was 13.44±0.98 which was gradually decreased and finally at day 90 it was reduced to 1.89±3.68. In NS group the mean Push tool total at baseline was 13.50±0.92 which was gradually decreased and finally at day 90 it was reduced to 4.61±4.37. Significant differences were found in the mean Push tool total between the PRP and NS groups on days 30 (p=0.015), 45 (p=0.016), 60 (p=0.003), and 75 (p=0.002) (Figure2). In the PRP and NS groups, the mean change in push tool total was significant from baseline to subsequent follow up periods (p<0.001) (Figure3).

**Figure1:**
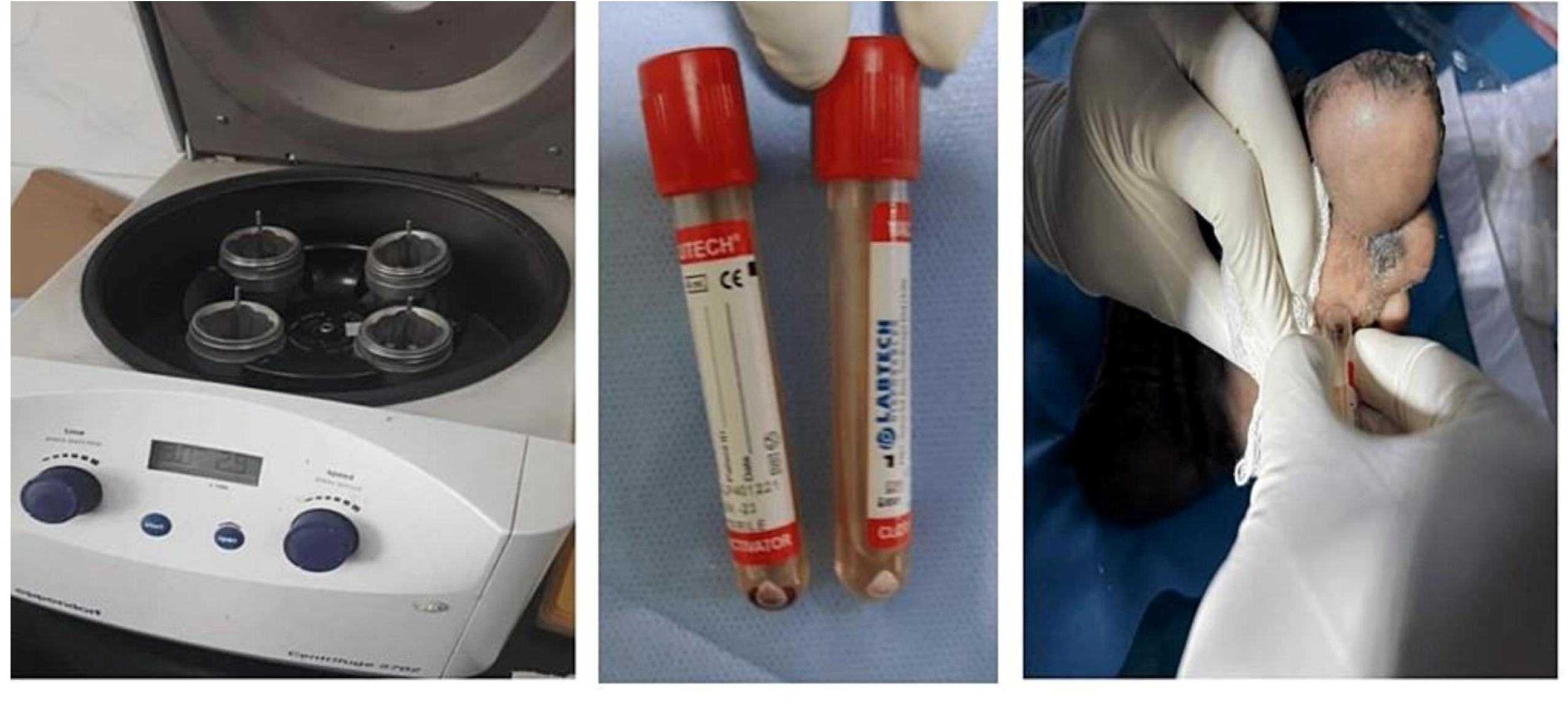
PRP Centrifugal machine (Eppendorf Centrifuge 5720), PRPandits application.

**Table–1:** Intergroup Comparison of Time to Heal.

| **Table–1: Intergroup Comparison of Time to Heal** |  |  |  |  |
| --- | --- | --- | --- | --- |
| Intervention | Time to heal (weeks) |  | Unpaired test t-value | p-value |
|  | Mean | SD |  |  |
| PRP group | 11.17 | 2.73 | -3.467 | 0.001 |
| NS group | 11.78 | 1.66 |  |  |

**Table–2:** Intergroup Comparison of Wound Size.

| **Table-2: Intergroup Comparison of Wound Size** |  |  |  |  |  |  |
| --- | --- | --- | --- | --- | --- | --- |
| Wound Size | PRP group |  | NS group |  | t-value | p-value |
|  | Mean | SD | Mean | SD |  |  |
| baseline | 8.28 | 1.18 | 8.45 | 1.13 | -0.44 | 0.665 |
| D15 | 7.70 | 1.17 | 8.19 | 1.28 | -1.19 | 0.243 |
| D30 | 5.92 | 1.58 | 7.76 | 1.46 | -3.62 | 0.001 |
| D45 | 4.43 | 1.75 | 6.40 | 1.40 | -3.73 | 0.001 |
| D60 | 2.83 | 1.78 | 4.86 | 1.38 | -3.82 | 0.001 |
| D75 | 1.32 | 1.79 | 3.24 | 1.60 | -3.40 | 0.002 |
| D90 | 0.61 | 1.20 | 1.58 | 1.55 | -2.10 | 0.044 |

**Table–3:**
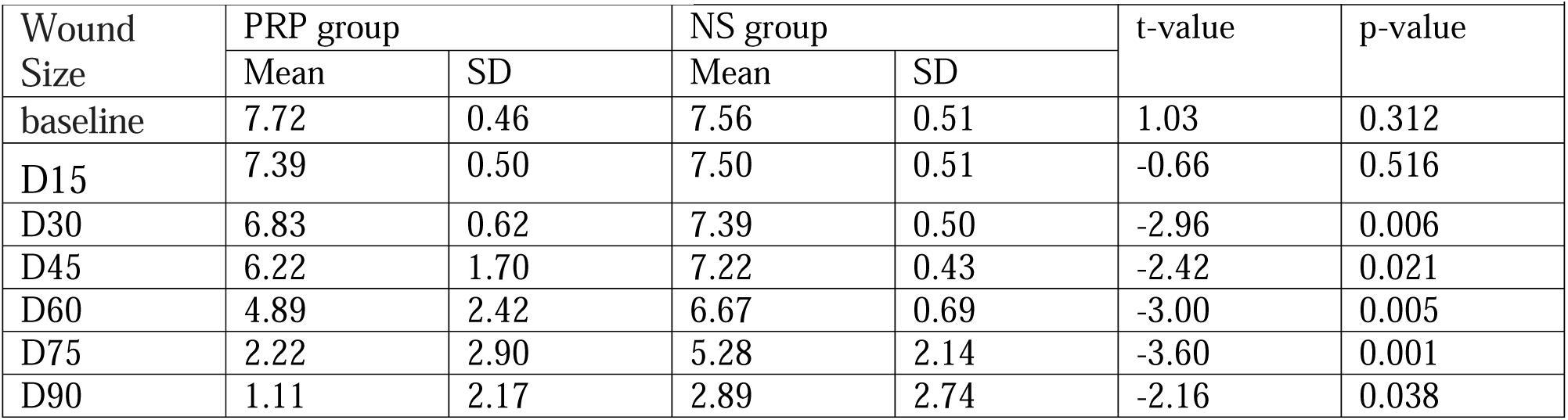
Intragroup Comparison of Wound Size.

| **Table–3: Intragroup Comparison of Wound Size** |  |  |  |  |  |  |
| --- | --- | --- | --- | --- | --- | --- |
| Wound Size | PRP group |  | NS group |  | t-value | p-value |
|  | Mean | SD | Mean | SD |  |  |
| baseline | 7.72 | 0.46 | 7.56 | 0.51 | 1.03 | 0.312 |
| D15 | 7.39 | 0.50 | 7.50 | 0.51 | -0.66 | 0.516 |
| D30 | 6.83 | 0.62 | 7.39 | 0.50 | -2.96 | 0.006 |
| D45 | 6.22 | 1.70 | 7.22 | 0.43 | -2.42 | 0.021 |
| D60 | 4.89 | 2.42 | 6.67 | 0.69 | -3.00 | 0.005 |
| D75 | 2.22 | 2.90 | 5.28 | 2.14 | -3.60 | 0.001 |
| D90 | 1.11 | 2.17 | 2.89 | 2.74 | -2.16 | 0.038 |

**Figure 2.**
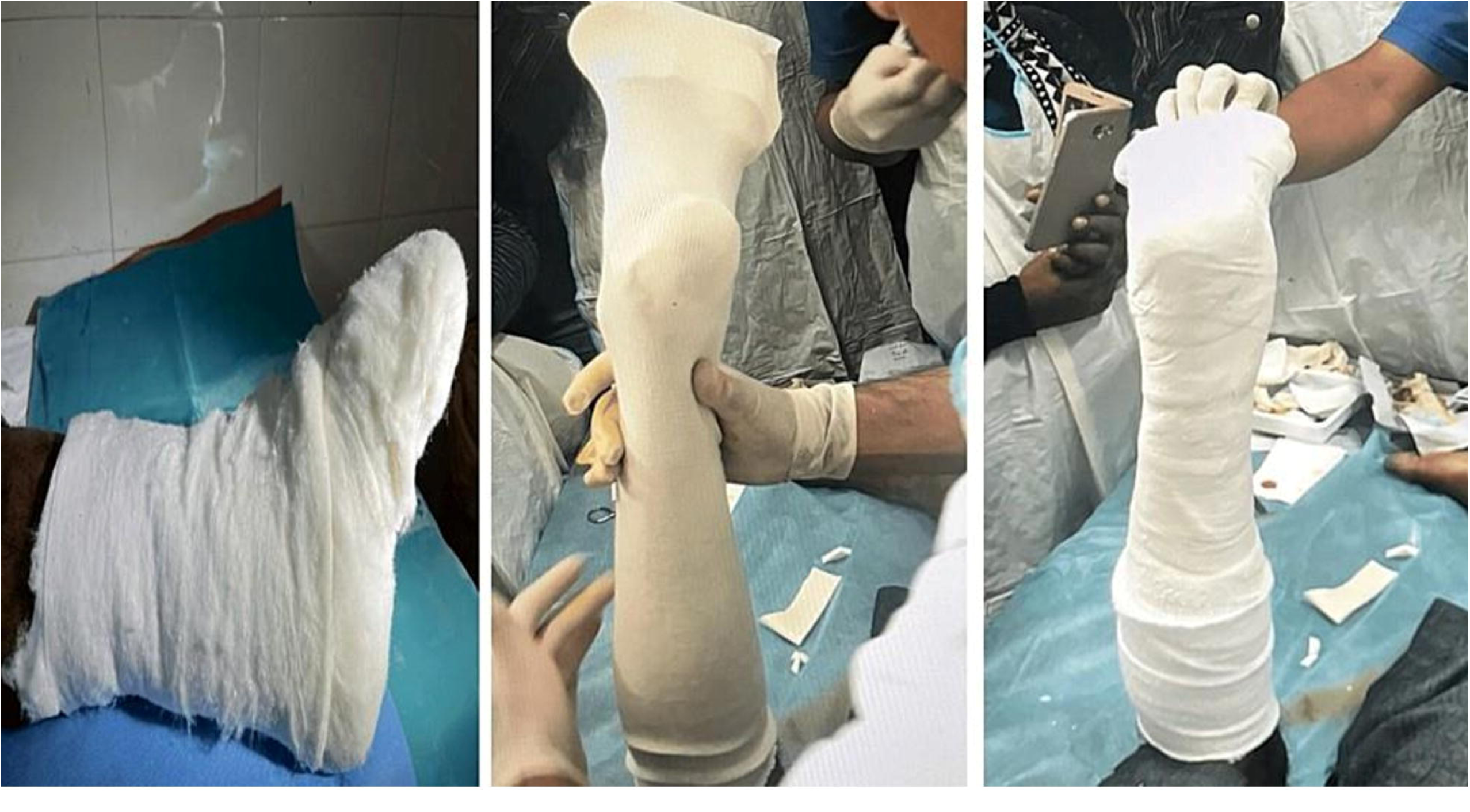
TCC application method.

**FIGURE3:**
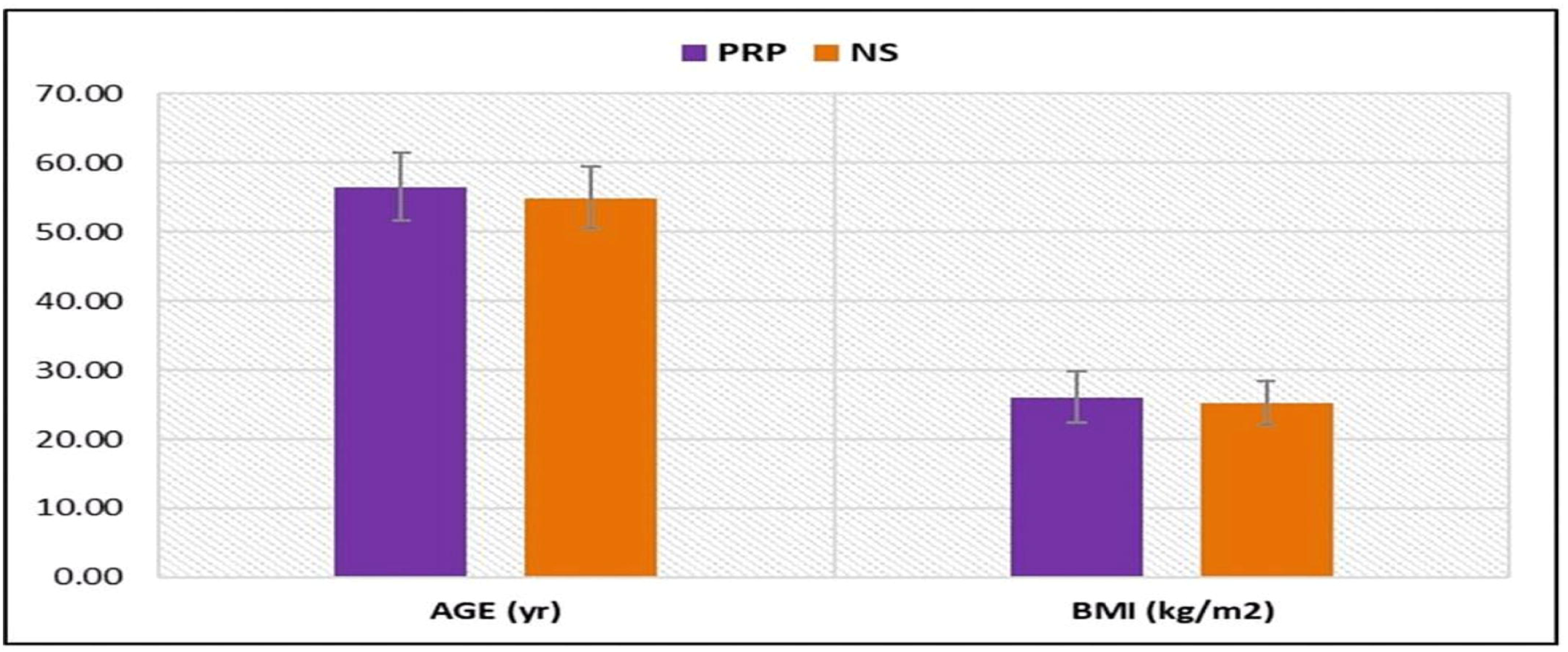
Distribution of Age & BMI in the groups.

## Discussion

Diabetic foot ulcers are a major health concern that can lead to arthopathy and limb amputation [16]. Microangiopathy and macroangiopathy modify the vascular status and increase the risk of infection due to hyperglycemia; peripheral neuropathy in the diabetic foot results in excessive foot pressure, foot deformities, and unstable gait. Foot care and glycemic index management are crucial for preventing the development of foot ulcers [17]. Identification of the “at-risk” foot, treatment of the acutely diseased foot, and prevention of further complications are the three main strategies for managing diabetic foot disease to avoid amputation of the lower extremities [18]. In many cases, aggressive treatment of DFUs can prevent worsening of the condition and the potential need for amputation [19]. Therefore, the objective of therapy should be early intervention to permit prompt healing of the lesion and once it has healed, to prevent its recurrence [20]. Platelet-rich plasma (PRP) is a portion of the plasma fraction of autologous blood with a platelet concentration above the baseline [21]. PRP is a growth factor agonist with mitogenic and chemo-tactic qualities that promotes wound healing via granules, which release locally acting growth factors [22, 23]. These growth factors promote tissue repair by attracting undifferentiated cells to the newly formed matrix and stimulating cell division [24]. Similar to the methods employed by Motolese et al., Shan et al., and Margolis et al., we used PRP as a gel dressing for DFU [25–27]. The gel was selected for practical use owing to its painless administration and patient preference. Moreover, compared to PRP injections into the floor and margins of the ulcer, the gel form might have a lower risk of infection. These factors increased the suitability of PRP gels for outpatient use. Since our institution uses normal saline as the standard dressing for DFU that are not infected, we decided to use it as the control group. In contrast to alternative therapeutic modalities that, while more successful, may have restricted access and high costs, especially in resource-constrained regions, this one is, after all, inexpensive, accessible, and easily available. Elsaid et al. found a similar proportion of sex (p=0.68), mean age, and BMI in the PRP group and NS, which were not significant (p=0.74) [28]. In this study, we found no statistical difference between the PRP and NS groups in terms of sex, size of the affected foot, age, BMI, duration of diabetes, ABI, RBS, HbA1C, serum albumin, or platelet count. Pressure reduction is an essential component in the treatment of diabetic foot ulcers. Total contact cast allows for mobility and relieves pressure over the ulcer, and has established the edit self as the gold standard for therapy, making patient adherence easier [29]. It has been shown to not only interrupt the pathogenesis chain that leads to ulceration but also induce changes in the histology of the ulcer [30]. The possible disadvantages of TCC include the need for specialized knowledge in its application, frequent cast replacements, and associated expenses [31]. According to Gupta et al; the mean duration of DM in the PRP group was 9.1±5.1 and in the NS group was 10.96±5.42, which was not statistically significant [32].

Ahmed et al. found that the mean ABI in the PRP group was 0.85±0.04, and that in the NS group was 0.83±0.01 with no statistically significant difference (p=0.881) [33]. Elsaid et al; (2019) and Ahmad et al; (2017) used a sample with statistically insignificant differences in HbA1c and mean RBS between the groups. The mean platelet count of the PRP group was 258.44±45.30, while that of the NS group was 243.22±42.32. No significant difference was found in the mean platelet count between the PRP and NS group (p=0.305), which is consistent with the finding of Elsaid et al; (2019) and Ahmed et al; (2017), where p values were 0.78 and 0.42 respectively. Elsaid et al; (2019) also compared the mean serum albumin levels between the PRP and NS groups, finding that they were 4.15±0.26 and 4.21±0.42, respectively (p=0.68). Additionally, they discovered that the PRP group’s mean duration to maximum healing was 4 weeks shorter than that of the control group (6.3±2.1vs.10.4± 1.7 weeks, P<0.0001). Likewise, Driver et al. showed that the PRP group recovered 28 days faster on average than the control group [34]. These results indicate that PRP’s proliferative effect may improve DFU healing. A significant difference was found in the mean time to healing between the PRP and NS group (p=0.001). Although it was statistically significant between the groups, it took much more time than the study by Elsaid et al; which may be due to fewer applications of PRP and keeping the wounds closed by TCC for 2weeks and wound inspection was not possible. In the PRP group, the total number of ulcers healed at 90 days was 14 out of 18 (77.78%), which was significantly better than that in the NS group (8 out of 18 ulcers, (44.44%) (p<0.05). This finding also correlates with the study by Elsaid et al; however, in contrast with Gupta et al; this difference may be due to differences in follow-up timings and different methods of application. When specific PUSH variables were analyzed separately, only the length × width decreased significantly among the healed ulcers. This may be due to the prevalence of stage 2 ulcers in the sample, the limited number of study ulcers with exudates, and the small number of categories used to distinguish changes in tissue type and exudate quantity. Gardner et al; (2005) found that the tissue type and exudate amount did not vary significantly at each follow-up, the only variable affecting the PUSH score was the tool’s length × width [35]. These parameters are consistent with the result of this study.

## Conclusion

In this study, we demonstrated that both PRP and NS dressings are beneficial and effective in the treatment of diabetic foot ulcers when used in conjunction with TCC to improve the healing rate. Although both treatments are beneficial, the results of this study indicated that PRP therapy is statistically superior to NS dressing. These results are consistent with those of the previous studies. The limitation of this study was the relatively short follow-up period. Diabetic foot ulcers can be chronic and slow healing wounds, often requiring extended periods for complete healing and for assessing the long-term efficacy of treatments. Therefore, a longer follow-up period would provide more comprehensive insights into the sustained effects of PRP therapy and NS dressing on diabetic foot ulcer healing rates.

**FIGURE4:**
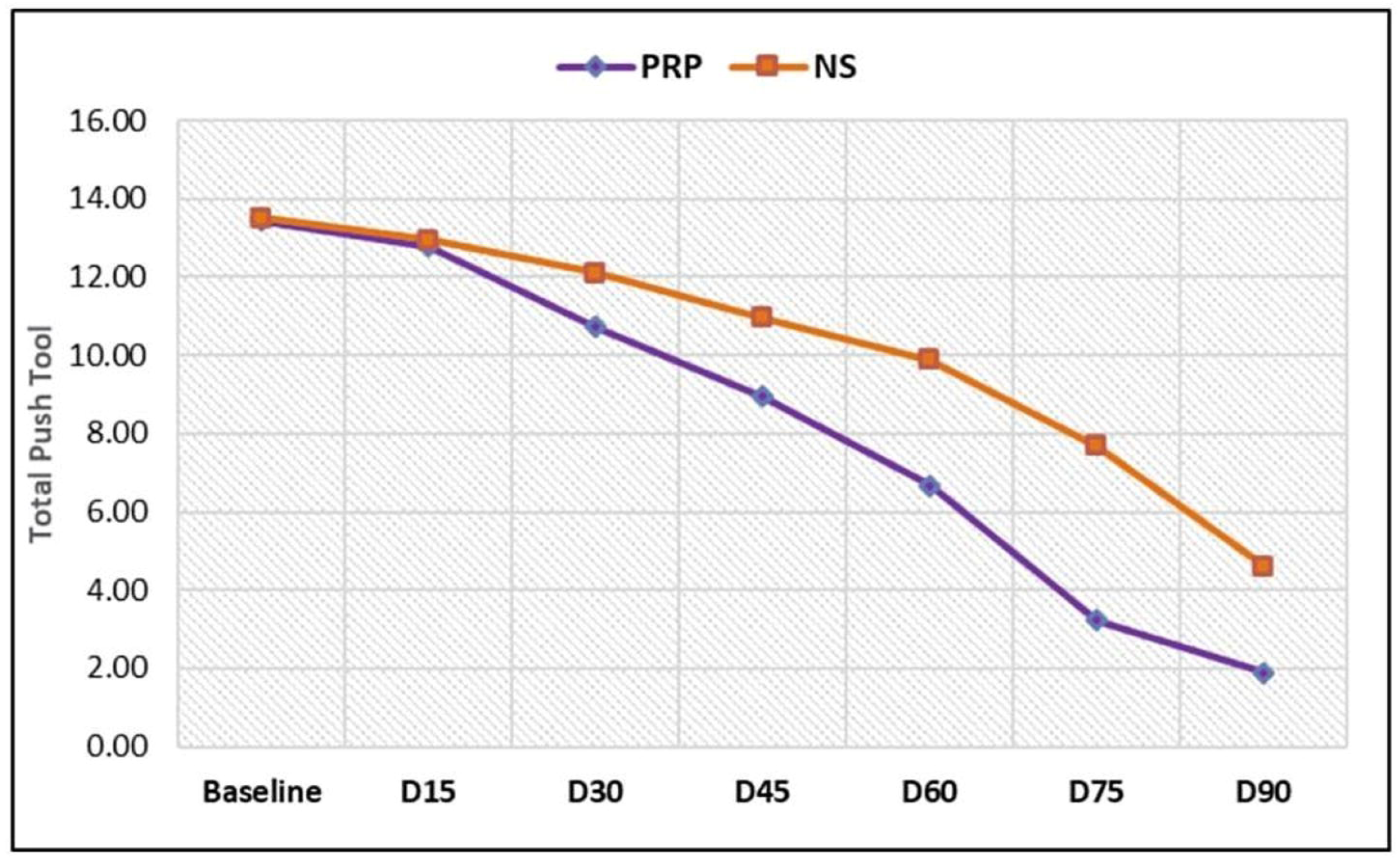
Push score total between PRP and NS groups.

**FIGURE5:**
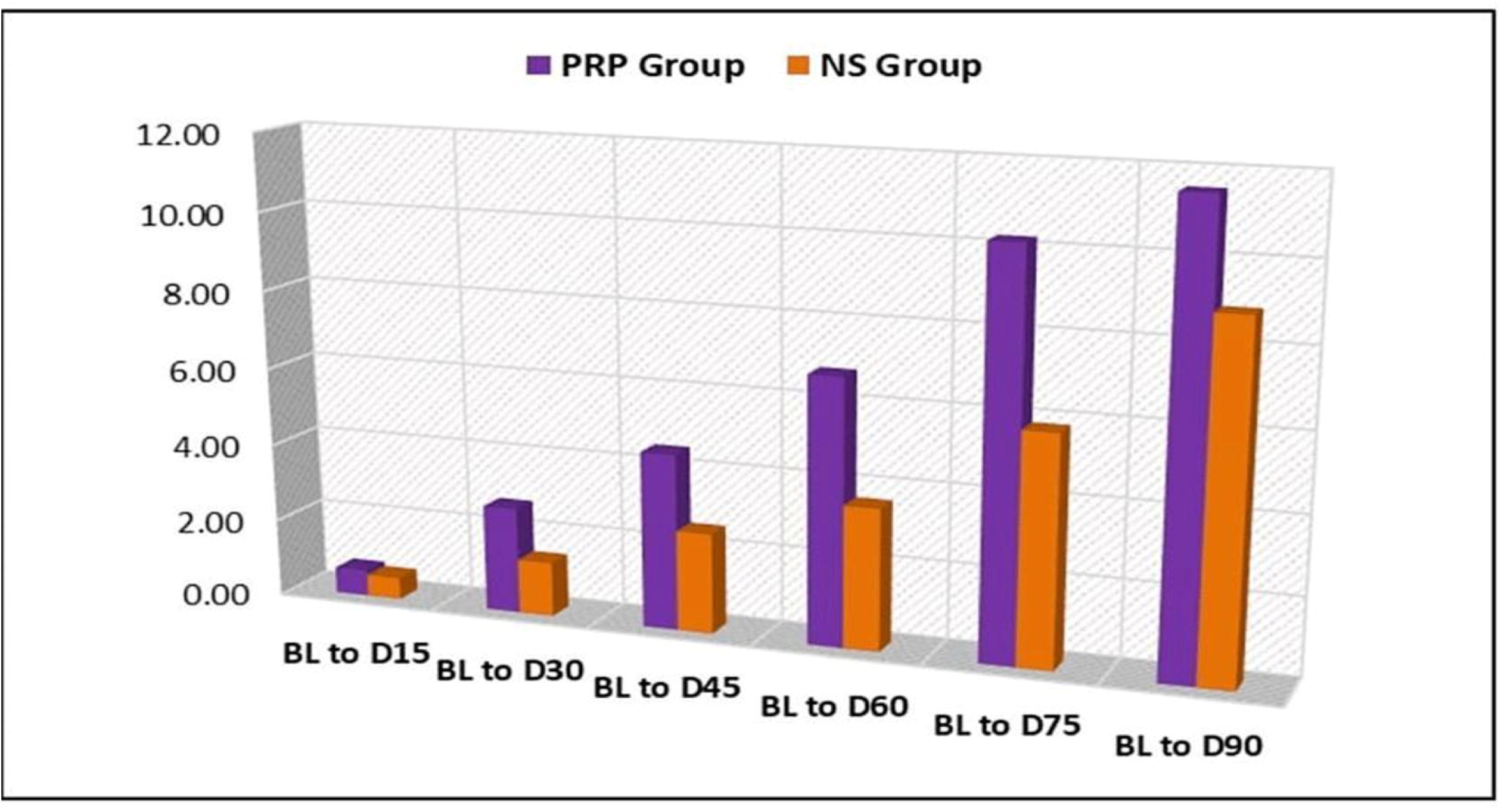
Intragroup Comparison of Push tool total between PRP and NS groups.

## Data Availability

Data collected from department of PMR AIIMS Patna.

## AVAILABILITY OF DATA

Data collected from department of PMR AIIMS Patna.

## Data Access Statement

All relevant data are within the paper and its Supporting Information files.

## Author Contribution

SD, NK contributed to the design and implemented research. SK, RK to the analysis and to the writing of the manuscript. SKP, AK conceived the original and supervised the project.

---

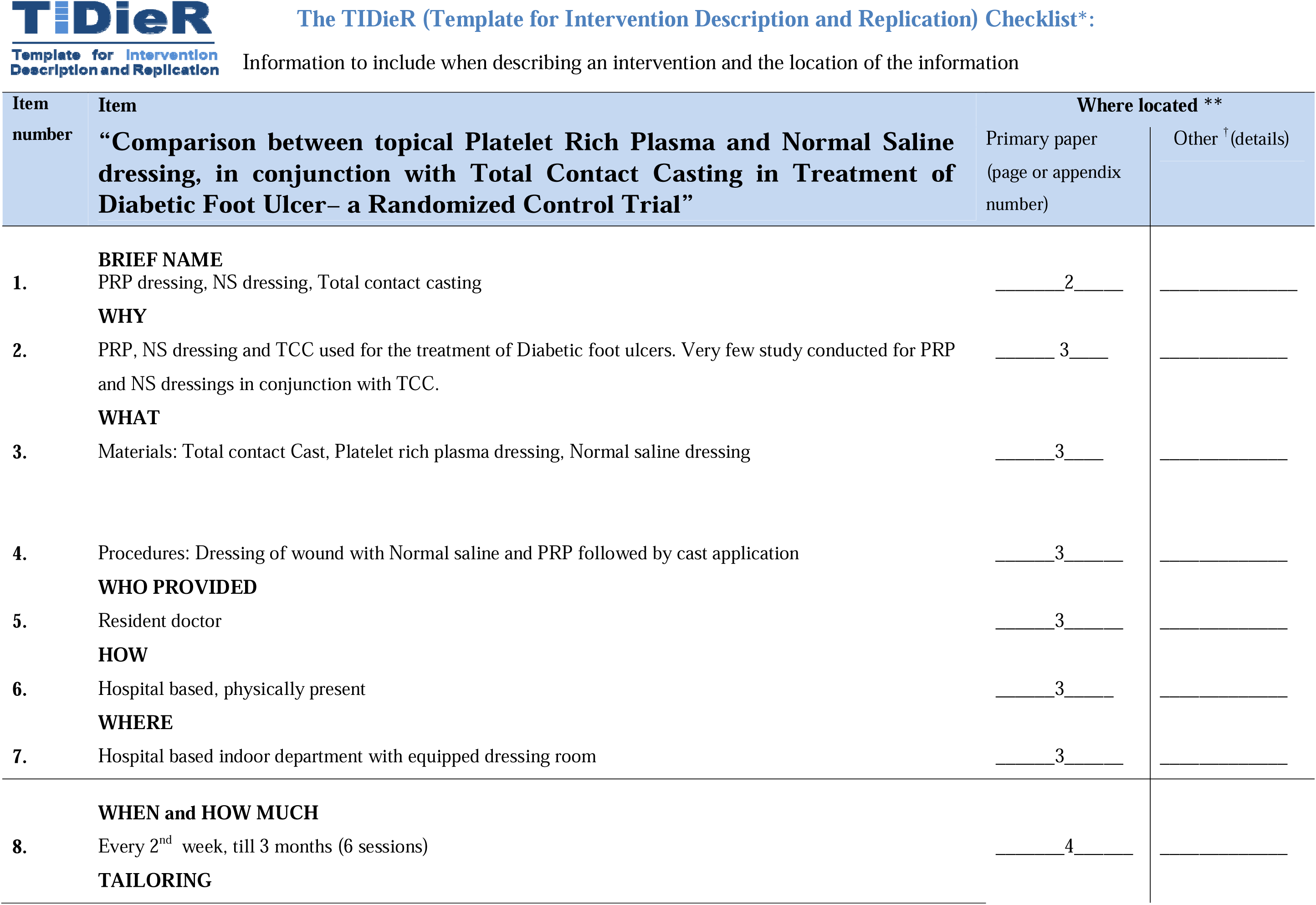
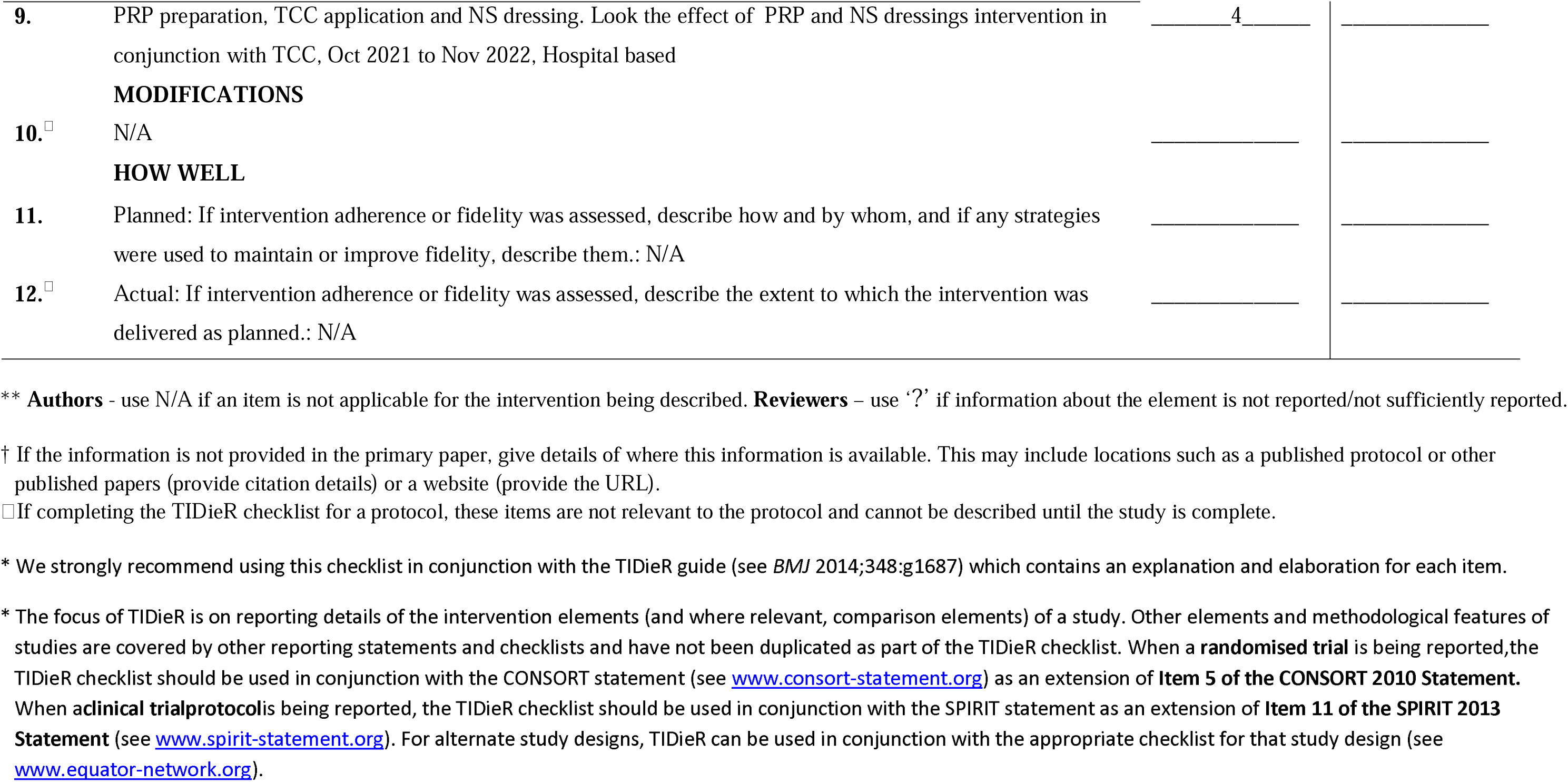

